# Threat of shock increases distractor susceptibility during the short-term maintenance of visual information

**DOI:** 10.1101/2023.11.22.23298914

**Authors:** Abigail Casalvera, Madeline Goodwin, Kevin Lynch, Marta Teferi, Milan Patel, Christian Grillon, Monique Ernst, Nicholas L Balderston

## Abstract

**BACKGROUND:** Work on anxiety related attention control deficits suggests that elevated arousal impacts the ability to filter out distractors. To test this, we designed a task to look at distractor suppression during periods of threat. We administered trials of a visual short-term memory (VSTM) task, during periods of unpredictable threat, and hypothesized that threat would impair performance during trials where subjects were required to filter out large numbers of distractors.

**METHOD:** Experiment 1 involved fifteen healthy participants who completed one study visit. They performed four runs of a VSTM task comprising 32 trials each. Participants were presented with an arrow indicating left or right, followed by an array of squares. They were instructed to remember the target side and disregard the distractors on the off-target side. A subsequent target square was shown, and participants indicated whether it matched one of the previously presented target squares. The trial conditions included 50% matches and 50% mismatches, with an equal distribution of left and right targets. The number of target and distractor squares varied systematically, with high (4 squares) and low (2 squares) target and distractor conditions. Trials alternated between periods of safety and threat, with startle responses recorded using electromyography (EMG) following white noise presentations.

Experiment 2 involved twenty-seven healthy participants who completed the same VSTM task inside an MRI scanner during a single study visit. The procedure mirrored that of Experiment 1, except for the absence of white noise presentations.

**RESULTS:** For Experiment 1, subjects showed significantly larger startle responses during threat compared to safe period, supporting the validity of the threat manipulation. However, results suggested that the white noise probes interfered with performance. For Experiment 2, we found that both accuracy was affected by threat, such that distractor load negatively impacted accuracy only in the threat condition.

**CONCLUSION:** Overall, these findings suggest that threat affects distractor susceptibility during the short-term maintenance of visual information. The presence of threat makes it more difficult to filter out distracting information. We believe that this is related to hyperarousal of parietal cortex, which has been observed during unpredictable threat.

## Introduction

Anxiety is the most commonly diagnosed class of mental health disorders among Americans^1^. Notably, anxiety significantly impairs individuals’ cognitive functioning and attentional abilities^2^. Impaired attention control and elevated arousal are key symptoms that cut across anxiety disorders^2,3^, but the degree to which these symptoms reflect common underlying mechanism is poorly understood.

One approach to studying the effects of arousal on attention control is to manipulate arousal during threat of unpredictable shock^4,5^. In this paradigm, subjects complete a cognitive task during periods of safety or unpredictable shock threat. During the threat periods, they know that the shock may come but the timing and frequency of the shocks is unpredictable. Changes in arousal are typically measured by quantifying the magnitude of the acoustic startle reflex^5,6^, elicited by a loud white noise. Increases in arousal potentiate this response, thus yielding a direct physiological measure of arousal changes during this experimental arousal induction paradigm^5,7–9^. Additionally, this instructed threat paradigm requires very few cognitive resources, thus making it possible to complete concurrent cognitive tasks during the threat periods ^10,11^. These details make the unpredictable threat paradigm a robust and ecologically valid approach to studying anxiety as well as anxiety cognition interactions. For this reason, this paradigm is widely used and previously shown to induce state anxiety, affect task performance, and alter patterns of functional connectivity^12–23^.

Previous research into the mechanisms mediating the threat related increases in arousal have implicated both cortical and subcortical regions like the amygdala, bed nucleus of the stria terminalis (BNST), insula, thalamus, and parietal cortex (x). Critically, work from our group has previously shown that regions of the parietal cortex exhibit both hyperactivity and hyperconnectivity during threat periods compared to safe periods^24,25^. It is well known that the parietal regions are critical for both endogenous and exogenous shifts of attention, suggesting that the effects of anxiety on attention control may be critically mediated by parietal cortex involvement ^26–29^. For example, the intraparietal sulcus is a region that is highly interconnected with the visual cortex, and known to be involved in attentional orienting ^30–32^. Accordingly, anxiety related hyperactivity of this region may increase orienting responses to external stimuli, resulting in an increase in vigilance to environmental stimuli ^30,31^. While adaptive evolutionarily, too much vigilance can increase distraction susceptibility and interfere with attention control ^33^.

To test the effects of arousal on attention control, we designed a visual short-term memory (VSTM) task where subjects were instructed to simultaneously encode (targets) and filter out (distractors) shapes in a visual array. Critically, the number of targets and distractors could be independently manipulated on each trial, allowing us to independently assess visual short-term memory encoding and distractor suppression across trials. Additionally, subjects performed this task during safe and (unpredictable) threat blocks, allowing us to assess the effect of arousal on each of these manipulations (target encoding, distractor suppression). In this study, we validated this task in a laboratory session where arousal was measured with the acoustic startle reflex (Experiment 1). We then had subjects complete the task in the MRI while we recorded BOLD activity. Given that anxiety is known to impair attention control, including the ability to ignore irrelevant information^34–37^, we hypothesized that anxiety-related attention control deficits might manifest as difficulties in selectively filtering out distractors during the threat condition. In addition, assuming parietal regions are involved in attention control, we hypothesized that threat related parietal activity may be related to performance on the task.

## Materials and Methods

### Participants

#### Experiment 1

Sixteen healthy volunteers from the Washington DC area were recruited for this experiment. One participant withdrew, leaving 15 participants who completed the session (7 female; *M* (SD): 29.47 (7.52) yo).

#### Experiment 2

Thirty-five healthy volunteers from the Washington DC area were recruited for this experiment. Four participants were excluded due to scheduling issues, and four participants were excluded due to scanner issues, leaving 27 participants who completed the session (13 female, *M* (SD): 30.33 (9.06) yo).

In both studies, participants were excluded if they had current or history of any axis I psychiatric disorder as assessed by SCID-I/NP, family history of mania, schizophrenia, or other psychoses, current or history of any psychotropic or illicit drug use. Contra-indications for fMRI, hearing loss, or any other medical conditions that might interfere with the study. All participants gave written informed consent approved by the NIMH Combined Neuroscience Institutional Review Board and received financial compensation.

### General Procedure

#### Experiment 1

Subjects arrived at the lab, completed the consent process, were briefed on the task, and were prepped for the procedure ^38^. The skin under the left eye was cleaned and prepped with an exfoliant gel, and afterward two electrodes were affixed directly below the left eye. Two additional electrodes were attached to the fingers of the right hand to measure skin conductance and serve as the ground for the EMG electrodes. Finally, two electrodes were attached to the left wrist to deliver the shock. Next the subject completed a shock workup procedure, which was used to calibrate the intensity of the shock. Afterward, they completed a white noise habituation run. Finally, they completed 4 runs of the VSTM task.

#### Experiment 2

The procedure for Experiment 2 was similar to Experiment 1, with the exception that subjects completed the task in the MRI scanner and did not receive white noise presentations. Subjects arrived at the lab, completed the consent process, were briefed on the task, and were prepped for the procedure. All metal was removed from the subjects’ person and the subject was given ear plugs for the scans. Two electrodes were attached to the left wrist to deliver the shock. Next the subject completed a shock workup procedure, which was used to calibrate the intensity of the shock. Finally, they completed 4 runs of the VSTM task.

### Materials

#### VSTM Task

On each trial, subjects were shown an arrow (150 ms) that pointed to either the left or right side of the screen. Subjects were instructed to attend to the side of the screen corresponding to the arrow direction and ignore the contralateral side of the screen. After a delay (2000 - 5000 ms), subjects were then shown an array of squares (150 ms) and instructed to remember the squares on the target side and ignore the squares on the off-target side. After another delay (2000 - 5000 ms), subjects were shown a single target square ipsilateral to the arrow direction and instructed to indicate whether or not the square matched one of the previously presented target squares. Half of the trials were matches; half were mismatches. Similarly, half of the trials targeted the left half of the screen; half targeted the right. The number of target and distractor squares was systematically varied across trials to include orthogonal high (4 squares) and low (2 squares) target and distractor conditions. Trial duration was 13 seconds, and the timing of the events within each trial was jittered. There was a total of 4 ∼8-minute runs, each with 32 scored trials and 3 dummy (shock) trials. Trials took place during alternating periods of safety and threat. During the safe period, subjects were not at risk of receiving a shock. During the threat periods, subjects were at risk for receiving a shock at any point during the entire threat period. There were 3 shocks per run. During Experiment 1, white noise presentations were presented during the retention intervals following square array presentations.

#### Acoustic Startle Stimulus

A 40-ms burst of a 103 dB white noise (near instantaneous rise/fall times) was delivered via the computer soundcard using standard over-the-ear headphones. Prior to the task, subjects completed a habituation block with 9 un-signaled presentations of the white noise.

#### Electromyography

Blink responses were measured using EMG recorded from the orbicularis oculi muscle with two tin cup electrodes placed under the left-eye. EMG was sampled at a rate of 2000 Hz using a Biopac data acquisition unit (MP150; Biopac Systems Inc., US) with Acknowledge 4.4 software.

#### Shock

A 100 ms, 200 Hz stimulation train was used as the aversive shock. The shock was delivered to the left wrist via 11mm disposable Ag/AgCl electrodes (EL508, Biopac Systems Inc., US), using a constant current stimulator (DS7A, Digitimer, LLC, Ft. Lauderdale, FL). Shock intensity was set prior to the experiment using a workup procedure, where subjects received a series of shocks of increasing intensity until they rated the stimulation as “uncomfortable but not painful”. This intensity was used for the remainder of the experiment.

#### EMG processing

EMG responses from Experiment 1 were analyzed using the analyze startle packaged developed by Dr. Balderston (https://github.com/balders2/analyze_startle). bandpass filtered (30–500 Hz), rectified, and smoothed with a 20-ms time constant. Blinks were scored as the peak during the response period (20–120 ms after white noise) minus the mean EMG signal during the baseline period (50 ms prior to white noise). Trials where no blink was detected (i.e. peak – mean was less than the range of the baseline) were scored as a 0. Trials where excessive noise was detected during the baseline (i.e. SD was greater than 2x the SD of the entire run) were scored as missing data ^38–42^. Blinks were then converted to t-scores ((X – M_X_)/SD_X_ * 10 + 50) to reduce large inter-individual differences in blink magnitude ^43^.

#### Scans

Scans were collected on a Siemens 3T Skyra MRI scanner with a 32-channel head coil. Subjects viewed the stimuli via a coil mounted mirror system. We acquired a T1-weighted MPRAGE (TR = 2530 ms; TE1 = 1.69 ms; TE2 = 3.55 ms; TE3 = 5.41 ms; TE4 = 7.27 ms; flip angle = 7°) with 176, 1 mm axial slices (matrix = 256 mm × 256 mm; field of view (FOV) = 204.8 mm × 204.8 mm). For functional data, we acquired whole-brain multi-echo echoplanar images (EPI; TR = 2000 ms; TEs = 13.8, 31.2, 48.6 ms; flip angle = 70°) with 32, 3 mm axial slices (matrix = 64 mm × 64 mm; FOV = 192 mm × 192 mm) aligned to the AC-PC line. We also acquired a reverse phase-encoded “blip” EPI image to correct for EPI distortion in the phase encoding direction.

#### fMRI Pre-processing

Data was processed using afni_proc.py ^44^, which included slice-timing correction, despiking, volume registration, identification of non-BOLD components using a TE-dependent independent components analysis (ICA) ^44^, scaling, EPI distortion correction, nonlinear normalization to the MNI template, and blurring with a 6 mm FWHM gaussian kernel. fMRI timeseries were scrubbed for motion (threshold set at >.5 mm RMS), and modeled using a first-level GLM. This GLM included regressors of no interest corresponding to the following: baseline (polynomial estimates), 6 motion parameters and their derivatives, the non-BOLD component timeseries, shock onsets, and button presses. The GLM also included regressors of interest corresponding to the cue presentation, the array presentation, and response prompt of the VSTM task.

### Statistical analysis

#### Startle

Startle data were processed and converted to t-scores. These scores were then analyzed using a paired sample t-test contrasting responses during safe and threat blocks.

#### Performance

Percent correct and average reaction time was calculated for each individual and each condition. No response trials were counted as missing data for reaction time scores. Accuracy and reaction time scores were analyzed using a 2 (condition: safe vs. threat) x 2 (target load: low vs. high) x 2 (distractor load: low vs. high) repeated measures ANOVA. Interaction effects were characterized by paired sample t-tests where appropriate. Partial eta-squared and Cohen’s d were calculated for ANOVA effects and t-tests, respectively.

#### fMRI analysis

The resulting beta maps from the first level GLM were then analyzed using a whole brain voxelwise analyses. We extracted the betas from the first level GLM corresponding to the cue and the array and performed mixed-model ANOVAs on the values for each event type. For the cue events, we used a 2 (condition: safe vs. threat) x 2 (cue direction: left vs. right hemisphere) ANOVA. For the array events we used a 2 (condition: safe vs. threat) x 2 (cue direction: left vs. right hemisphere) x 2 (target load: low vs. high) x 2 (distractor load: low vs. high) repeated measures ANOVA. We used cluster-based thresholding to correct for multiple comparisons by conducting 10,000 *Monte Carlo* simulations ^45^. We used a 2-tailed voxelwise p-value of 0.001, and a non-Gaussian autocorrelation function that better approximates BOLD data to estimate the smoothness of the residuals ^46^, and clustered voxels with adjoining faces and edges. The result was a minimum cluster size of 55, 3 mm isotropic voxels.

## Results

### Experiment 1

#### Startle

As a manipulation check, we examined the effect of threat on startle responses. As designed, subjects showed significantly larger startle responses during threat compared to safe periods (t(14) = 2.82; p = 0.014; d = 0.73).

#### Accuracy

To determine the effect of threat on VSTM performance, we performed a 2 (condition: safe vs. threat) x 2 (target load: low vs. high) x 2 (distractor load: low vs. high) repeated measures ANOVA on accuracy scores. We found a significant main effect of target load (f(1,14) = 20.61; p = 0; eta-squared = 0.6), suggesting that subjects were more accurate on low load compared to high load trials. We found no other significant main effects or interactions (all ps > 0.1).

#### Reaction time

Like accuracy scores we performed a 2 (condition: safe vs. threat) x 2 (target load: low vs. high) x 2 (distractor load: low vs. high) repeated measures ANOVA on reaction time. Like accuracy, we found a significant main effect of target load (f(1,14) = 19.54; p = 0.001; eta-squared = 0.58), suggesting that subjects were faster on the low compared to high load trials. In addition, we found a significant condition by target load by distractor load interaction (f(1,14) = 6.92; p = 0.02; eta-squared = 0.33; all other ps > 0.1). However, when we analyzed the safe and threat trials separately with 2 (target load: low vs. high) x 2 (distractor load: low vs. high) repeated measures ANOVAs, we found no differential effects for safe trials compared to threat trials (all ps > 0.1). The only effects observed were significant main effects for target load for both the safe (f(1,14) = 10.61; p = 0.006; eta-squared = 0.43) and threat (f(1,14) = 22.21; p = 0; eta-squared = 0.61) trials, consistent with the main effect from the 2x2x2 ANOVA. Although we didn’t directly test this hypothesis, we believe that some of this variability is due to interference of the startle probes on performance.

### Experiment 2

#### Accuracy

As with Experiment 1, we performed a 2 (condition: safe vs. threat) x 2 (target load: low vs. high) x 2 (distractor load: low vs. high) repeated measures ANOVA on accuracy scores. For accuracy, we found a significant main effect for both condition (f(1,26) = 10.61; p = 0.003; eta-squared = 0.29) and target load (f(1,26) = 33.18; p = 0; eta-squared = 0.56). We also found a trend toward a main effect for distractor load (f(1,26) = 3.56; p = 0.071; eta-squared = 0.12), which was likely driven by a significant condition by distractor interaction (f(1,26) = 5.72; p = 0.024; eta-squared = 0.18; all other ps < 0.1).

When we analyzed accuracy for safe and threat trials separately, we found distinct patterns for the safe and threat trials. For safe trials, we found a significant main effect for target load (f(1,26) = 38.22; p = 0; eta-squared = 0.6), but no main effect for distractor load (f(1,26) = 0.16; p = 0.691; eta-squared = 0.01) and no target load by distractor load interaction (f(1,26) = 0.03; p = 0.856; eta-squared = 0). For threat trials, we found significant main effects for both target load (f(1,26) = 10.41; p = 0.003; eta-squared = 0.29) and distractor load (f(1,26) = 9.58; p = 0.005; eta-squared = 0.27), but no target load by distractor load interaction (f(1,26) = 0; p = 0.957; eta-squared = 0).

#### Reaction time

As with Experiment 1, we performed a 2 (condition: safe vs. threat) x 2 (target load: low vs. high) x 2 (distractor load: low vs. high) repeated measures ANOVA on reaction time. Although we found significant main effects for both target load (f(1,26) = 34.11; p = 0; eta-squared = 0.57) and distractor load (f(1,26) = 5.09; p = 0.033; eta-squared = 0.16), these main effects were likely driven by significant condition by target load (f(1,26) = 19.77; p = 0; eta-squared = 0.43) and condition by distractor load interactions (f(1,26) = 14.09; p = 0.001; eta-squared = 0.35; all other ps > 0.1).

As with accuracy, we found distinct patterns for the safe and threat trials. For safe trials, we found a significant main effect of distractor load (f(1,26) = 12.84; p = 0.001; eta-squared = 0.33), but no main effect for target load (f(1,26) = 0.35; p = 0.56; eta-squared = 0.01) and no condition by target load interaction (f(1,26) = 0.07; p = 0.798; eta-squared = 0). In contrast, for threat trials we found a significant main effect for target load (f(1,26) = 66.43; p = 0; eta-squared = 0.72), but no main effect for distractor load (f(1,26) = 1.83; p = 0.187; eta-squared = 0.07) and no condition by target load interaction (f(1,26) = 1.29; p = 0.266; eta-squared = 0.05).

#### fMRI Results

For both the cue and the array presentations, we primarily see visual evoked activity differentiating between trials where subjects were instructed to fixate on the left vs. the right hemisphere. For the cue presentations, this activity is greatest in the hemisphere contralateral to the focus of attention, replicating previous findings showing that top-down attentional instructions can bias visual processing. Additionally, for the array presentations, we se bilateral threat related activity in a pair of clusters that include voxels in both the striatum and the insula, which has also been previously shown to be responsive to threat. Finally, in parietal cortex, we see a threat by attentional focus interaction. During the safe period, we see greater activity when attention is focused on the left hemisphere. In contrast, during the threat period, we see greater activity when attention is focused on the right hemisphere.

## Discussion

In this study, we investigated the impact of threat of shock on attentional control and distractor suppression during a visual short-term memory task. Our research aimed to explore the interaction between anxiety-related arousal and attentional processes, building on previous findings related to anxiety and its effects on cognitive functioning. The findings give compelling evidence to support our initial hypothesis, that heightened anxiety-related arousal impairs visual short term memory performance when there are more distractors. In the safe conditions of our study, the presence of distractors slowed down participants’ reaction times without compromising their accuracy, indicating a typical pattern of attentional interference ^47,48^.

However, intriguingly, under the threat condition, there was no significant impact on reaction time, suggesting that heightened arousal did not influence the speed of processing information. Instead, the threat condition significantly affected accuracy, indicating that participants’ ability to filter out distractors and maintain precision in their responses was compromised ^49^. These findings suggest that threat-induced arousal does indeed affect distractor susceptibility, disrupting the cognitive processes responsible for selective attention. Furthermore, we suggest that the instructed threat blocks biased subjects toward more automatic (and less) accurate response patterns. Consistent with conclusion, previous literature suggests that heightened arousal impairs the cognitive mechanisms responsible for focusing attention and ignoring irrelevant stimuli^34,36^. Our neuroimaging data revealed distinct patterns of activity in the parietal cortex across threat conditions: during safe conditions, increased activity was observed on trials where attention was directed to the left hemifield, while under threat conditions, increased activity was observed on trials where attention was directed to the right hemifield. This interaction between threat conditions and attentional focus in the parietal cortex offers mechanistic support for the conclusion that threat affects distractor susceptibility, with the parietal cortex playing a central role in mediating this effect. These results underscore the intricate relationship between anxiety-induced arousal, attentional control, and the underlying neural mechanisms, shedding light on the complex interplay between cognitive processes and emotional states ^12^.

Participants engaged in effortful attentional processes during the safe trials to effectively filter out distractors, indicating a deliberate and controlled mechanism of attention. However, under the threat condition, a shift occurred in their attentional strategy. Participants seemed to rely on automatic processes, as reflected in the lack of significant changes in reaction time.

Paradoxically, this automatic mode of attentional processing during threat led to a notable decrease in accuracy, suggesting that the filtering process was impaired under heightened arousal. This shift in attentional mechanisms appeared to involve a lateralization effect: effortful attentional processes might have driven attention toward the left hemisphere, while automatic attentional processes predominantly activated the right hemisphere. This intriguing finding aligns with the existing literature on attentional processes, particularly in cases of spatial neglect, where individuals tend to ignore stimuli presented on one side ^50,51^. Building upon this model, a testable hypothesis emerges, and future experiments could investigate the neural underpinnings of this attentional switch, potentially exploring the involvement of specific regions within the parietal cortex and their connectivity patterns. This study’s results are consistent with previous research, providing valuable insights into the complex interplay between attention, arousal, and hemispheric specialization, laying the foundation for further explorations in the field of cognitive neuroscience (x).

These findings hold significant relevance in the context of anxiety, shedding light on how heightened anxiety impacts the ability to filter out distractors and respond to threats effectively. The results suggest that anxiety might disrupt the balance between deliberate attentional control and automatic, stimulus-driven processes ^52,53^. Under threat, automatic distractor suppression seems to be bolstered, indicating a heightened vigilance towards potentially threatening stimuli. However, this increased vigilance comes at a cost, and the accuracy of responses suffer. In the face of anxiety, individuals may find themselves more prone to automatic, reflexive responses ^54^, prioritizing the rapid identification and suppression of potential threats over the careful, effortful evaluation of task-related information. This shift in attentional dynamics not only highlights the intricacies of anxiety-related attention deficits but also underscores the multifaceted challenges faced by individuals experiencing heightened anxiety. Understanding these processes is crucial, not only for advancing our knowledge of the neural mechanisms underlying anxiety but also for developing targeted interventions that can help individuals regulate their attentional responses in anxiety-provoking situations, ultimately improving their cognitive functioning and overall well-being.

The significant increase in participants’ startle responses under threat conditions serves as an indicator of the successful induction of anxiety, validating the experiment’s ability to create an anxious state ^55^. Moreover, our study effectively tests attentional control by examining the differential activation patterns in the visual cortex during the cue and square array presentation phases. Notably, the convergence of startling stimuli and the threat condition in our study is of great significance. It activated key brain regions associated with emotional processing, such as the insula and thalamus, emphasizing the emotional intensity of the induced anxiety. Interestingly, the striatum, a region traditionally associated with motivation, also featured prominently in our findings ^56^. While the exact role of the striatum in this context warrants further exploration, its involvement suggests a potential link between anxiety-induced arousal and motor responses, hinting at the complex interconnections within the brain’s neural circuitry ^56,57^. This analysis provides valuable insights into the dynamic shifts in attentional focus and highlights the intricate neural processes involved in selective attention under varying emotional states. The experiment’s validity is further reinforced by the design, allowing for the exploration of the underlying neural mechanisms governing attentional regulation. By systematically manipulating the threat level and number of target or distractor squares, our study provides a comprehensive understanding of the intricate interplay between anxiety, attentional control, and threat processing.

### Strengths and Limitations

Our study’s strengths lie in the within-subject anxiety manipulation, enhancing result reliability. Fear induction through the threat of shock adheres to established practices, providing a robust anxiety-inducing method. Utilizing diverse measurements, including behavior, physiology (startle responses), and neuroimaging (fMRI), ensures a comprehensive understanding of anxiety’s impact on attention. The dual-experiment design and transparent open science practices enhance internal validity and contribute clinically relevant insights for anxiety disorders.

Interpreting our findings is contingent upon acknowledging several limitations. The relatively small sample size may constrain the generalizability of our results, especially across diverse populations, given our primary recruitment focus on the Washington D.C. area. Furthermore, our study exclusively examined the immediate effects of anxiety-induced arousal, offering insights into immediate responses but overlooking the enduring, chronic nature of attentional deficits in anxiety-related disorders. The absence of a sham or control condition in our experimental design poses a significant limitation, as it complicates distinguishing the effects attributed to the threat of shock from those influenced by potential confounding variables. Incorporating a sham or control condition in future studies would bolster the research’s rigor, providing a more robust basis for evaluating the impact of arousal-induced anxiety on attentional processes.

## Conclusions

In summary, this investigation illuminates the intricate dynamics between heightened anxiety, attentional control, and cognitive functions. Our findings suggest a shift from effortful to automatic attentional processes under threat, manifested by sustained reaction times but diminished accuracy. The observed lateralization effect in the parietal cortex underscores the nuanced interplay between hemispheric specialization and anxiety’s impact on attention. Looking ahead, this study prompts further exploration with an emphasis on larger, more diverse samples, specific brain region investigations, and the inclusion of control conditions. These future endeavors are pivotal for refining our comprehension of anxiety-related attentional mechanisms. This research not only deepens our understanding of anxiety’s influence on attention but also lays the groundwork for nuanced explorations of cognitive processes within emotional contexts, offering potential avenues for targeted interventions and enhanced neural models.

## Author contributions

CRediT author statement according to: https://www.elsevier.com/authors/policies-and-guidelines/credit-author-statement.

**Conceptualization:** CG, ME, NLB

**Methodology:** CG, NLB

**Software:** NLB

**Formal analysis:** KGL, NLB

**Investigation:** MG, NLB

**Writing - Original Draft:** AC, NLB

**Writing - Review & Editing**: AC, MG, KGL, MT, MP, CG, ME, NLB

**Visualization:** AC, NLB Supervision: CG, ME, NLB

**Project administration:** CG, NLB

**Funding acquisition:** CG, NLB

**Figure 1.**
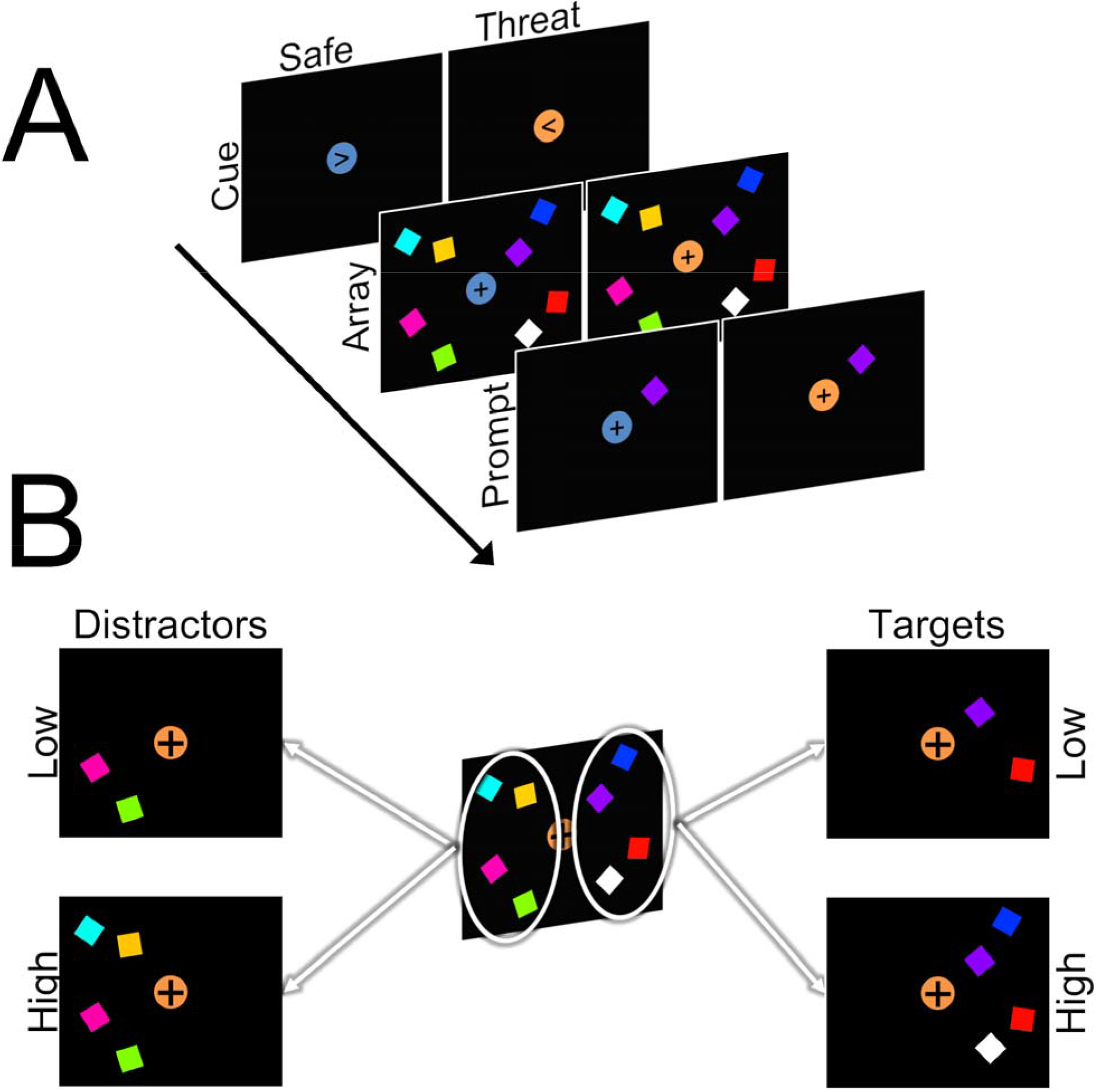
Schematic of task design. **A)** Trials began with a cue (150 ms) period where a centrally presented arrow pointed either to the left or right. Following a short delay, an array of target (ipsilateral to arrow direction) and distractor (contralateral to arrow description) squares were presented (150 ms). Following another delay, a single prompt square was presented (1000 ms). Subjects were instructed to indicate whether or not the prompt square matched one of the previous target squares. B) The number of target and distractor squares was systematically varied across trials to include orthogonal high (4 squares) and low (2 squares) target and distractor conditions.

**Figure 2.**
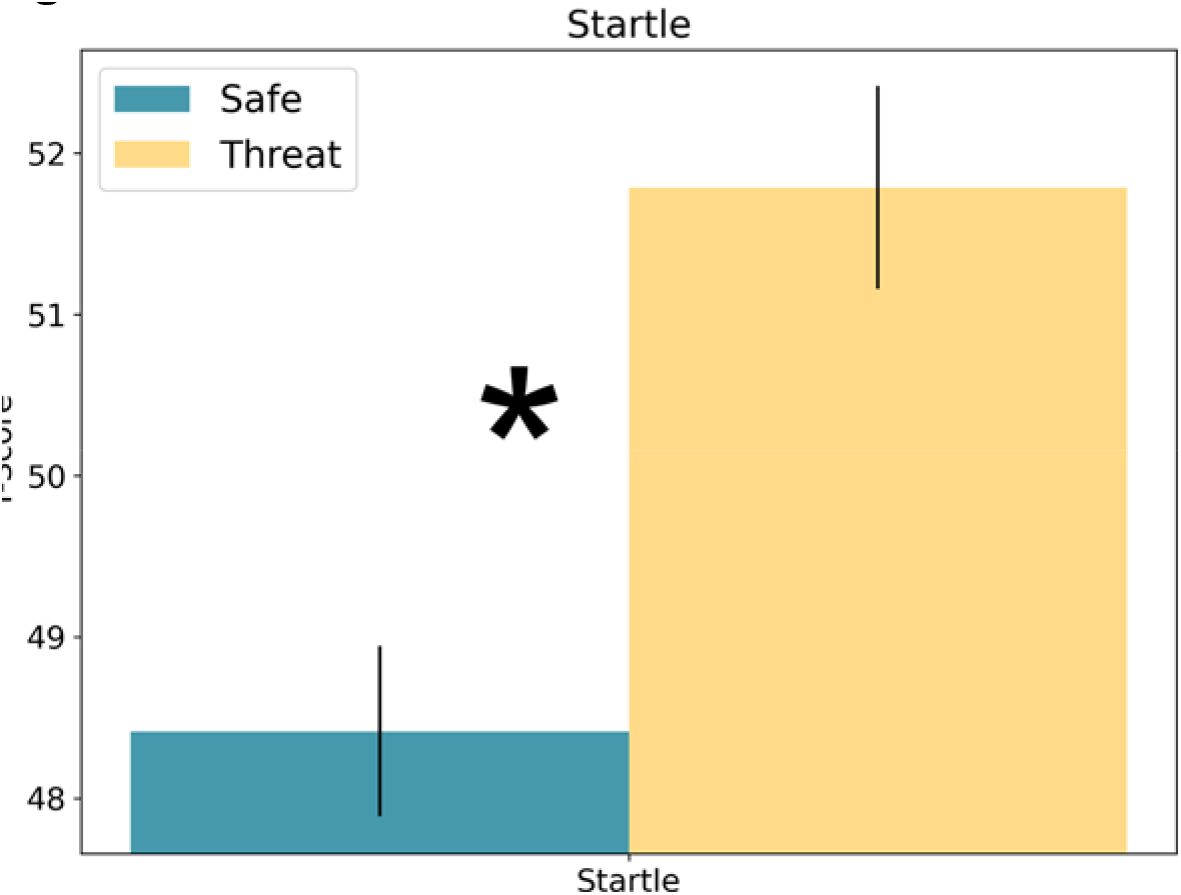
Startle responses during Experiment 1. Raw startle responses were converted to T-scores (X - M_X_)/SD_X_ *10 + 50). Bars represent the mean ± SEM. * = p < 0.05.

**Figure 3.**
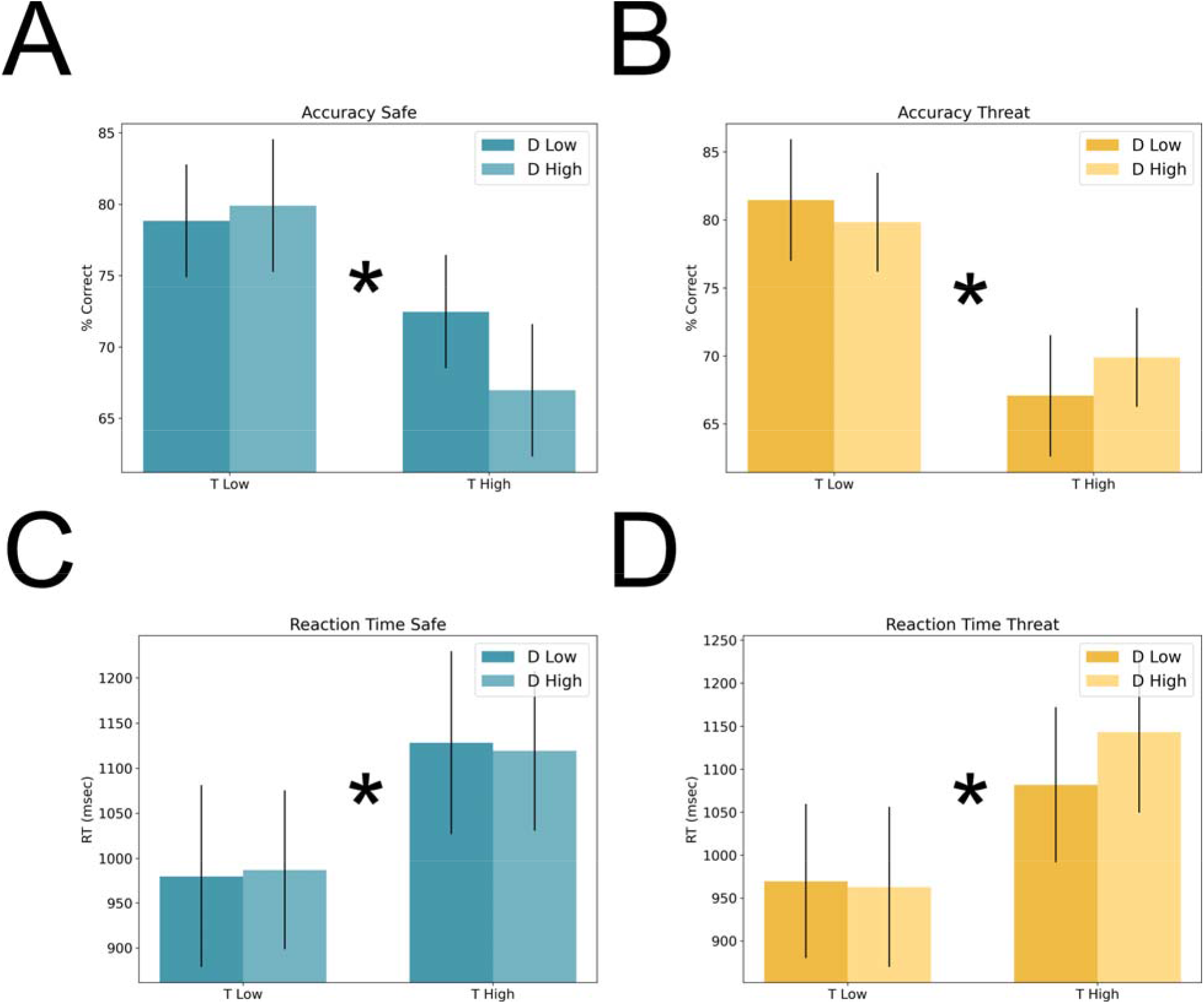
Accuracy and reaction time during Experiment 1. **A)** Percent correct during the safe periods of the VSTM task. **B)** Percent correct during the threat periods of the VSTM task. **C)** Reaction time during the safe periods of the VSTM task. **D)** Reaction time during the threat periods of the VSTM task. Bars represent the mean ± SEM. * = p < 0.05

**Figure 4.**
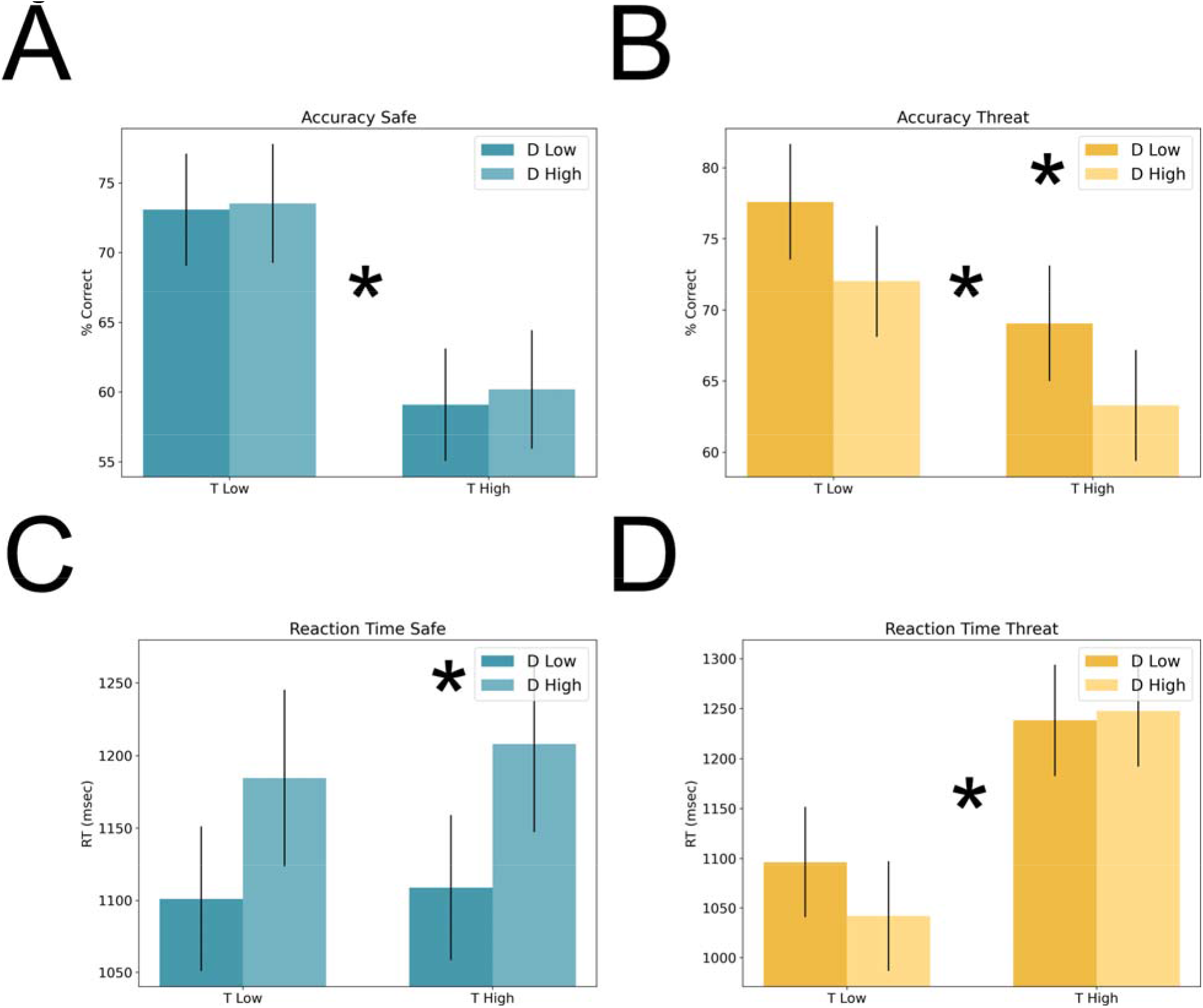
Accuracy and reaction time during Experiment 2. **A)** Percent correct during the safe periods of the VSTM task. **B)** Percent correct during the threat periods of the VSTM task. **C)** Reaction time during the safe periods of the VSTM task. **D)** Reaction time during the threat periods of the VSTM task. Bars represent the mean ± SEM. * = p < 0.05

## Data Availability

All data produced in the present study are available upon reasonable request to the authors

## Acknowledgments

The study team would like to thank the following individuals who contributed to Dr. Balderston’s K01 project: Dr. Kerry Ressler, Dr. Michael Thase, and Dr. Kristin Linn. The study team would like to also thank the DSMB members who oversaw the project: Dr. Lindsay Oberman (Chair), Dr. Alex Shackman, Dr. Gang Chen. This study utilized the high-performance computational capabilities of the CUBIC computing cluster at the University of Pennsylvania. (https://www.med.upenn.edu/cbica/cubic.html). The authors would like to thank Maria Prociuk for her expertise and assistance in submitting the paper. We would also like to thank the participants for their time and effort.

## Ethical Standards

The authors assert that all procedures contributing to this work comply with the ethical standards of the relevant national and institutional committees on human experimentation and with the Helsinki Declaration of 1975, as revised in 2008.

